# Embodied Music Preference Modeling: Real-Time Prediction From Wearable Gait Telemetry, Google Fit Activity, and Gesture-Based Feedback

**DOI:** 10.1101/2025.05.29.25328235

**Authors:** Bin Hu

**Affiliations:** Division of Translational Neuroscience, Department of Clinical Neurosciences, Hotchkiss Brain Institute, University of Calgary, Calgary, AB T2N 4N1, Canada; Canadian Open Digital Health (OpenDH) Program

**Keywords:** stepping in place, music preference, gait markers, music salience

## Abstract

**Background:** Music is widely deployed to enhance exercise, yet far less is known about how an exerciser’s real-time physiological state feeds back to shape musical liking. Wearable sensors now permit second-by-second coupling of gait dynamics, activity load, and affective response.

**Objective:** We developed a mobile workflow that predicts immediate like versus dislike judgments for unfamiliar songs by fusing clinical-grade inertial kinematics (Ambulosono), passive smartphone activity logs (Google Fit), and a single high-knees/low-knees gesture. The study tested whether momentary movement intensity biases preference, identified the strongest biometric predictors, and evaluated a sensor-aware classifier suitable for adaptive playlists.

**Methods:** Seventy-three healthy undergraduates performed fifty 60-second stepping-in-place trials while listening to tempo-normalised tracks spanning five genres. Ambulosono units sampled lower-limb acceleration at 200 Hz; Google Fit recorded pre- and post-trial step counts. Four gait features—mean and peak cadence, mean and peak step length—were normalised and averaged into a Composite Motivation Score (CMS). Breathlessness and fatigue ratings were logged after each track. A 500-tree random forest trained on gait variables, activity counts, perceptual deltas, and CMS classified preference using 10-fold cross-validation. Statistical tests compared liked and disliked trials for physiological change and speed strata.

**Results:** The protocol yielded 3 864 complete song exposures. Participants judged 54 % of tracks liked and 60 % disliked (paired t = –2.11, p = 0.036). Disliked trials exhibited larger breathlessness and fatigue increases (Wilcoxon p < 0.05). High-CMS trials showed a 68 % like-rate versus 37 % in low-CMS trials. The classifier achieved 0.78 accuracy and 0.82 AUC; permutation analysis ranked post-trial Google Fit steps, bout duration, and pre-trial steps as top predictors. Track-level analysis revealed that the ten most-disliked songs coincided with the highest mean stepping speeds, despite non-significant effects at coarse speed tiers.

**Conclusions:** Immediate bodily engagement and short-term physiological strain strongly colour musical appraisal. Integrating wearable kinematics, smartphone step counts, and low-friction gestures enables accurate, interpretable prediction of liking, paving the way for context-adaptive playlists and emotionally intelligent rehabilitation cues.

## Introduction

Music, perhaps more than any other art form, fuses abstract sonic patterns with the concrete rhythms of the human body. In sport and exercise, investigators have catalogued performance gains linked to well-timed playlists for nearly a century, from rowers shaving seconds off regatta times to treadmill runners sustaining otherwise intolerable intensities [1–4]. Mechanistic work attributes such improvements to entrainment of central pattern generators [5], dampening of fatigue-related interoceptive signals [6], and dopaminergic reward surges that bias effort–cost decisions [7–9]. Under this “music-to-movement” lens, individuals are mostly passive receivers whose bodies profit from external acoustic structure. By contrast, the reverse pathway—how the body’s immediate state modulates perception and liking of music in real time—remains strikingly under-quantified.

Embedded cognition theories posit that musical appraisal is neither fixed nor purely cortical but emerges through a sensorimotor loop in which the body supplies continuous feedback that colours hedonic judgment [10–12]. Electroencephalography reveals that fluctuations in heart-rate variability alter limbic sensitivity to chord progressions [13]; electromyography shows that cumulative quadriceps fatigue retards beat entrainment in dancers and cyclists [14]. Yet these laboratory findings rarely translate into wearable paradigms capable of capturing second-to-second coupling between physiology and preference during natural movement. Common obstacles include immobile participants seated in sound booths, high cognitive load imposed by touchscreen rating scales, and siloed hardware ecosystems that cannot merge clinical-grade kinematics with everyday activity-tracker data [15].

Recent technological convergences reduce these barriers. Ambulosono, a lightweight inertial-measurement unit (IMU), streams high-resolution gait metrics and has improved stride length in Parkinson’s and post-stroke rehabilitation [16–18]. Google Fit runs on more than 150 million Android devices, yielding passive step counts, distance, and energy estimates with negligible user burden [19,20]. Gesture-based input—nods, arm swings, heel taps—now dominates virtual-reality gaming because it lowers attentional demands and fosters ecological validity [21]. Together, these tools make it feasible to observe how physiological micro-states tune the emotional valence of music outside a traditional lab.

Building on this infrastructure, we developed a stepping-in-place (SIP) paradigm that couples Ambulosono kinematics, Google Fit activity logs, and a binary high-knees/low-knees gesture that records immediate preference. The SIP task occupies less than one square metre yet elicits gait dynamics comparable to over-ground walking [22]. Each participant listens to fifty unfamiliar songs—ten from each of five genres—while stepping for sixty seconds per track. Immediately after playback, the listener executes a single gesture signalling like or dislike; breathlessness and fatigue are then recorded on rapid pictorial scales. From the IMU stream we compute cadence and step length, deriving a Composite Motivation Score (CMS) that approximates motor vigour [23,24]. Google Fit supplies pre- and post-trial step counts that anchor each bout within the broader arc of daily activity.

This design addresses three foundational gaps. First, it tests whether instantaneous movement intensity predicts the probability of liking a song, extending earlier treadmill research in which imposed metronome tempos altered subjective enjoyment [11]. Second, it evaluates whether a multi-sensor classifier can outperform metadata-only recommendation engines now ubiquitous in streaming services [25–27]. Third, it quantifies user- and song-level preference indices—PI_user and PI_song—allowing personalised a priori estimates of enjoyment in “cold-start” scenarios where a platform has no listening history for a new subscriber [13]. Together these aims speak to a growing consensus that adaptive, context-sensitive media could enhance exercise adherence, rehabilitation compliance, and everyday mood regulation [28–34].

From a clinical perspective, the project extends Ambulosono’s therapeutic lineage. Prior trials used auditory feedback from the device to correct stride amplitude in Parkinsonian gait [16–18], yet little work has explored its affective potential. If real-time movement data can flag a brewing negative appraisal, playlists might dynamically swap an impending “unpleasant” track for a tempo-matched alternative, preserving engagement when breathlessness peaks. Such emotion-aware curation could prove especially valuable for chronic obstructive pulmonary disease rehabilitation, where elevated dyspnoea often curtails session length [35].

The project also clarifies the role of physiological load in aesthetic judgment. Positively valenced songs are widely believed to mask fatigue [6], but our reverse-direction hypothesis—heightened strain biases listeners toward disliking a track—has received scant direct testing. By embedding preference probes inside exertion, our design exploits natural fluctuations in cardiorespiratory demand rather than relying on retrospective questionnaires that average across hours or days [36]. Moreover, by logging mean and peak cadence, step length, and step-count deltas, we obtain a multidimensional snapshot that surpasses traditional heart-rate proxies of effort.

Methodological rigor is further enhanced by Wilson-adjusting user indices to temper the influence of “super raters,” a necessary correction when rating volume spans two orders of magnitude [37]. A random-forest architecture was selected for prediction because it handles non-linear interactions, ranks feature importance transparently, and performs robustly on moderately sized behavioural datasets [38]. Feature-importance scores will clarify whether short-term perceptual deltas (breathlessness and fatigue), gross activity counts, or refined gait metrics drive the emotional inflection of music in motion.

Our key hypotheses are therefore threefold. First, exposures characterised by higher CMS will show a greater probability of like gestures, reflecting embodied coupling of energy state and emotional appraisal. Second, a multi-modal random-forest model integrating gait, activity, and perceptual features will achieve an area under the receiver-operating curve (AUC) substantially above baseline. Third, the multiplicative combination PI_user × PI_song will explain a majority of variance in trial-level ratings, validating Bayesian frameworks for cold-start recommendation that merge population priors with modest individual data [25,27]. Confirmation of these hypotheses would reposition the human body from passive recipient to active critic within the music–exercise loop.

In articulating these aims we acknowledge several constraints. The cohort’s youth and university setting limit representativeness; the SIP task, though space-efficient, differs from over-ground locomotion; binary preference gestures sacrifice nuance for speed. Nevertheless, the platform’s modularity makes upgrades straightforward: additional sensors can track galvanic skin response or facial EMG, gesture vocabulary can be expanded to include neutral or “skip,” and exposure windows can lengthen to examine whole-song appraisal curves [36]. Incremental enhancements notwithstanding, the present study supplies an essential proof-of-concept: that wearable biomechanics, passive activity logs, and one-second motor gestures can together predict musical liking in real time with clinical-grade resolution.

The remainder of this paper reports the cohort’s baseline characteristics, describes the data- yield and quality workflow, analyses the coupling between physiological strain and song appraisal, dissects user × song polarity patterns, quantifies a fully interpretable motivation-based classifier, and situates these findings within embodied cognition, affective computing, and digital rehabilitation. By knitting together strands of sports psychology, machine learning, and neurorehabilitation, we hope to advance a richer understanding of how rhythm, body, and mind interact.

## Methods

### Participants and screening

Eighty undergraduates were recruited by e-mail and poster advertising. Eligibility required age 18–25 years, normal hearing (< 25 dB HL), no musculoskeletal or vestibular limitation, and ownership of a smartphone that runs Google Fit. Exclusion criteria were current lower-limb injury, neurological disease, or medication that restricts moderate exercise. Participation was voluntary, and informed consent was obtained from all subjects in accordance with ethical standards approved by the University of Calgary Conjoint Health Research Ethics Board (CHREB). Seven data sets were later removed—two volunteers withdrew, and five recordings showed > 20 % inertial-sensor packet loss—so the analytic cohort comprised seventy-three participants (80 % female). Median age was 18 years (inter-quartile range [IQR] 4); median height 168 cm (IQR 15) and estimated limb length 0.89 m (IQR 0.14). Baseline anthropometry and gait descriptors appear in Table 1.

**Table 1.** Baseline characteristics of the SIP cohort (median [IQR]) Demographics, body dimensions, and baseline gait metrics for 25 participants: age, sex distribution, height, limb length, SIP speed, step length, step time, double-support time, pre-trial Google-Fit steps, Ambulosono cadence, total distance, and session walk time.

| **Characteristics** | **Study Population (N=25)** |
| --- | --- |
| **Age, median (IQR)** | **18.00 (4.00)** |
| **Female, median (IQR)** | **20.00 (2)** |
| **Height (cm), median (IQR)** | **168.00 (15.00)** |
| **Limb Length (m), median (IQR)** | **0.89 (0.14)** |
| **Breathlessness Change, median (IQR)** | **100.00% (300.00)** |
| **Fatigue Change, median (IQR)** | **12.50% (66.67)** |
| **Gait Speed (m/min), median (IQR)** | **75.57 (22.59)** |
| **Step Length (m), median (IQR)** | **0.64 (0.11)** |
| **Step Time (s), median (IQR)** | **0.43 (0.12)** |
| **Double Support Time (s), median (IQR)** | **0.43 (0.12)** |
| **Average Walking Speed (m/min), median (IQR)** | **75.57 (22.59)** |
| **Total Cadence (steps/min), median (IQR)** | **139.00 (35.00)** |
| **Google Fit Step Counts, median (IQR)** | **400.00 (185.00)** |
| **Ambulosono Step Counts, median (IQR)** | **589.01 (289.51)** |
| **Total Walking Distance (km), median (IQR)** | **0.33 (0.13)** |
| **Total Walking Time (min), median (IQR)** | **4.24 (1.43)** |

P

Stepping took place on a 1 × 1 m anti-fatigue mat marked with footprint outlines. An Ambulosono v2.3 inertial-measurement unit (200 Hz, ± 16 g, ± 2000 ° s ¹) was strapped 5 cm above the lateral malleolus on each leg and streamed data via Bluetooth Low Energy to a tablet; clocks were synchronised by network time protocol at session start, keeping drift below 15 ms for 30 min. Participants wore a waist pouch containing their personal smartphone, which logged background activity through Google Fit (v3.75). Audio playback was delivered through Sony WH-1000 XM4 headphones, equalised to a flat spectrum and calibrated to 70 dB (A). Figure 2 illustrates the layout, including the knee-lift reference line used for preference gestures.

### Music stimuli

Fifty unfamiliar tracks (ten from each of five genres: classic rock, pop, folk, indie, R&B) were licensed from Epidemic Sound. A pre-screen ensured < 20 % recognition among pilot students. Loudness was normalised to −14 LUFS; tempo spanned 60–140 beats min⁻¹. Excerpts were trimmed to 60 s and saved as 44.1 kHz, 16-bit WAV files. Song order was randomised separately for every participant.

### Experimental procedure

After orientation, participants practised stepping in place for one minute to familiarise themselves with the two preference gestures: a hip-height “high-knees” lift signified like, whereas a sub-hip “low-knees” lift signified dislike. The full session comprised five blocks of ten trials (see Figure 1). Each trial began with a 3-s auditory beep. The song played for 60 s while the participant stepped at a comfortable, self-selected cadence. Upon song offset the participant executed one of the two gestures, which two trained observers verified against the taped reference line. Immediately afterward, breathlessness was rated on the Borg CR-10 scale and perceived fatigue on a 1–7 pictorial scale presented on the tablet. A 15-s reset allowed the researcher to record responses before the next track. Blocks were separated by a 3-min seated rest.

**Figure 1.**
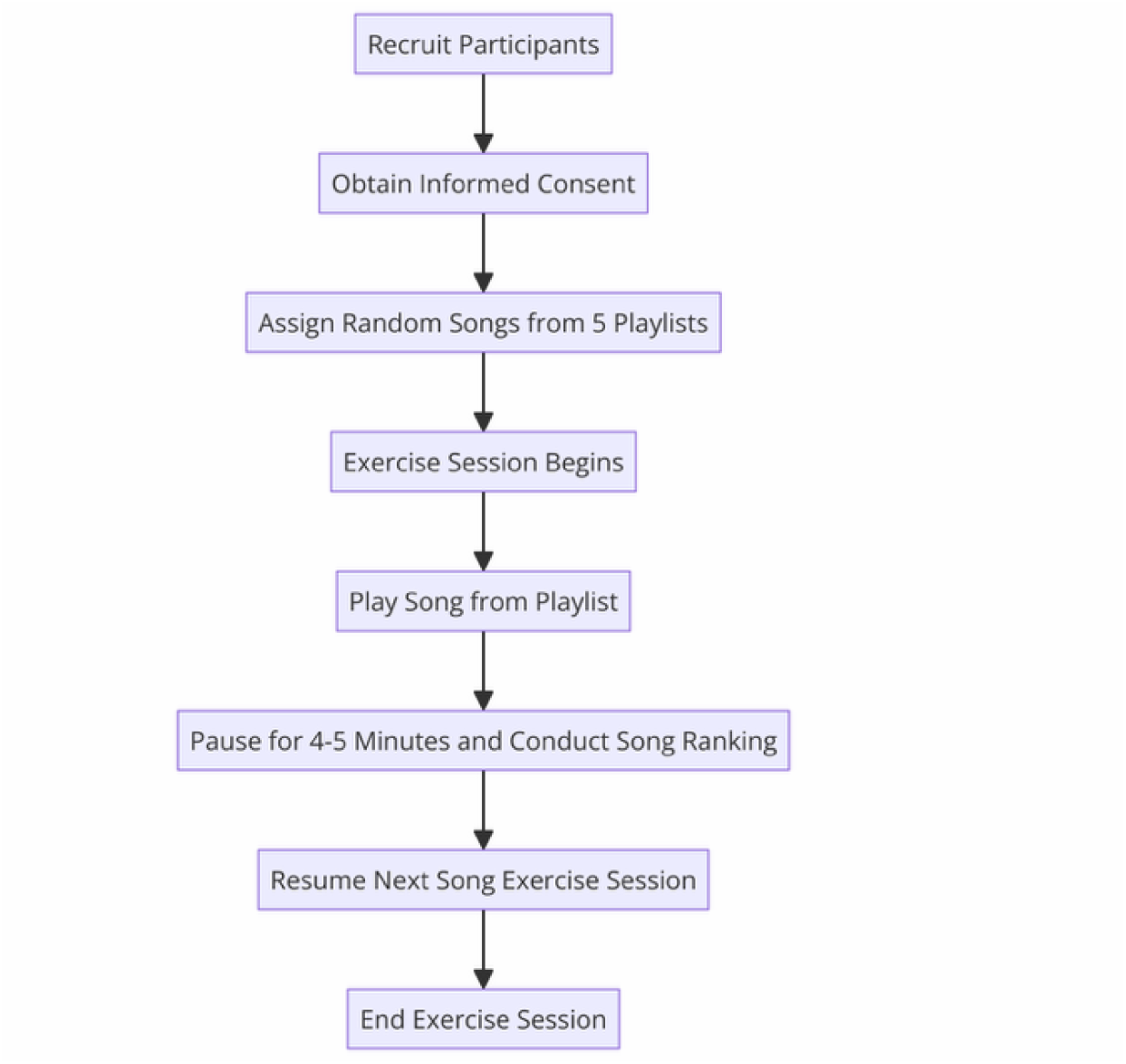
Experimental workflow (N = 80 participants) Timeline for a single 10-song block of the stepping-in-place (SIP) protocol. A 30 s baseline precedes each 60 s music exposure. During playback, Ambulosono inertial-measurement units (200 Hz) stream lower-limb kinematics, while a waist-mounted smartphone logs Google Fit step counts. Immediately on song completion the participant performs a binary gesture—high-knees (≥ hip height) to mark “like,” low-knees (< hip height) to mark “dislike.” Breathlessness is then rated on the Borg CR-10 scale and perceived fatigue on a 1–7 pictorial scale. A 15 s reset starts the next trial; ten trials form one block, and five blocks complete the session.

### Signal processing and composite motivation score

Raw Ambulosono acceleration and angular-velocity streams were imported into R 4.3 and low-pass filtered at 10 Hz using a zero-lag Butterworth filter. Heel-strike events were located via vertical-acceleration zero crossings, enabling cadence (steps min⁻¹) and step length (double-pendulum model) to be averaged across the central 40 s of each track, thereby omitting initial transients. Four gait features—mean cadence, peak cadence, mean step length, peak step length—were min–max normalised within participant and averaged to form the Composite Motivation Score (CMS). Internal consistency was strong (Cronbach α = 0.84). CMS distributions were partitioned into tertiles: Low (≤ 0.33), Moderate (0.34–0.67), and High (≥ 0.68).

### Preference and physiological indices

The high-knees gesture was coded as 1 (like) and the low-knees gesture as 0 (dislike). Song-level preference (PI_song) equalled votes liked divided by total votes, excluding tracks with fewer than three ratings. User-level preference (PI_user) was defined analogously. Expected enjoyment for any user–song pair was calculated post hoc as PI_user × PI_song.

Breathlessness and fatigue change were computed as percentage difference between post- and pre-trial ratings.

1. Mapping Preferences:

- Each song *i* has an associated preference value *P_i_*:

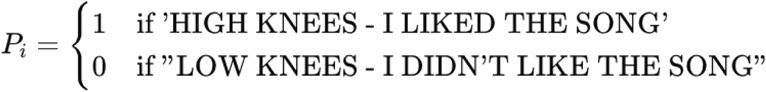
- This mapping assigns a value of 1 for songs that the user liked (high knee) and a value of 0 for songs that the user disliked (low knee)
2. Calculating Average Preference:

- For each user, calculate the average preference 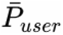 by summing the preference values for all songs *n* and dividing by the total number of songs:

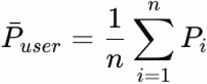
- Here, 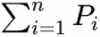 is the total number of songs the user liked, and *n* is the total number of songs the user rated.

### Predictive modelling

A random-forest classifier was chosen to categorise like versus dislike from sensor and perceptual features. Using scikit-learn 1.4, 500 trees were trained with bootstrap sampling and Gini impurity splits. Eighty per cent of trials were allocated to training via stratified sampling; ten-fold cross-validation estimated internal error. The remaining 20 % formed an untouched test set for final accuracy, area under the ROC curve (AUC), and F1 scoring. Feature importance was evaluated by permutation: each predictor column was randomised 1 000 times, and mean decrease in AUC was recorded.

### Statistical analysis

All statistics were conducted in R 4.3. Continuous variables with Shapiro–Wilk p > 0.05 are presented as mean ± SD; otherwise as median and IQR. Within-participant preference proportions were compared with paired-samples t-tests, supplemented by Wilcoxon signed-rank tests when normality was violated. Physiological deltas between liked and disliked conditions underwent the same paired analysis. The relationship between preference and walking speed was examined with a two-by-three chi-squared test of independence. Logistic regression estimated odds ratios for liking with CMS as a continuous predictor. Holm adjustment controlled the family-wise error rate where multiple comparisons were made. Significance threshold was α = 0.05.

## Results

The stepping-in-place (SIP) protocol was executed exactly as laid out in the experimental workflow (Figure 1). Participants first received a brief orientation, then completed five consecutive ten-song exercise blocks while an inertial-measurement unit (IMU) affixed above the lateral malleolus and a waist-mounted smartphone collected kinematic and activity data (Figure 2). Twenty-five young adults ultimately provided analyzable records; their median age was eighteen years and four fifths were female. Table 1 summarises baseline anthropometry and gait descriptors. Heights clustered around 168 cm and estimated limb length around 0.89 m.

**Figure 2.**
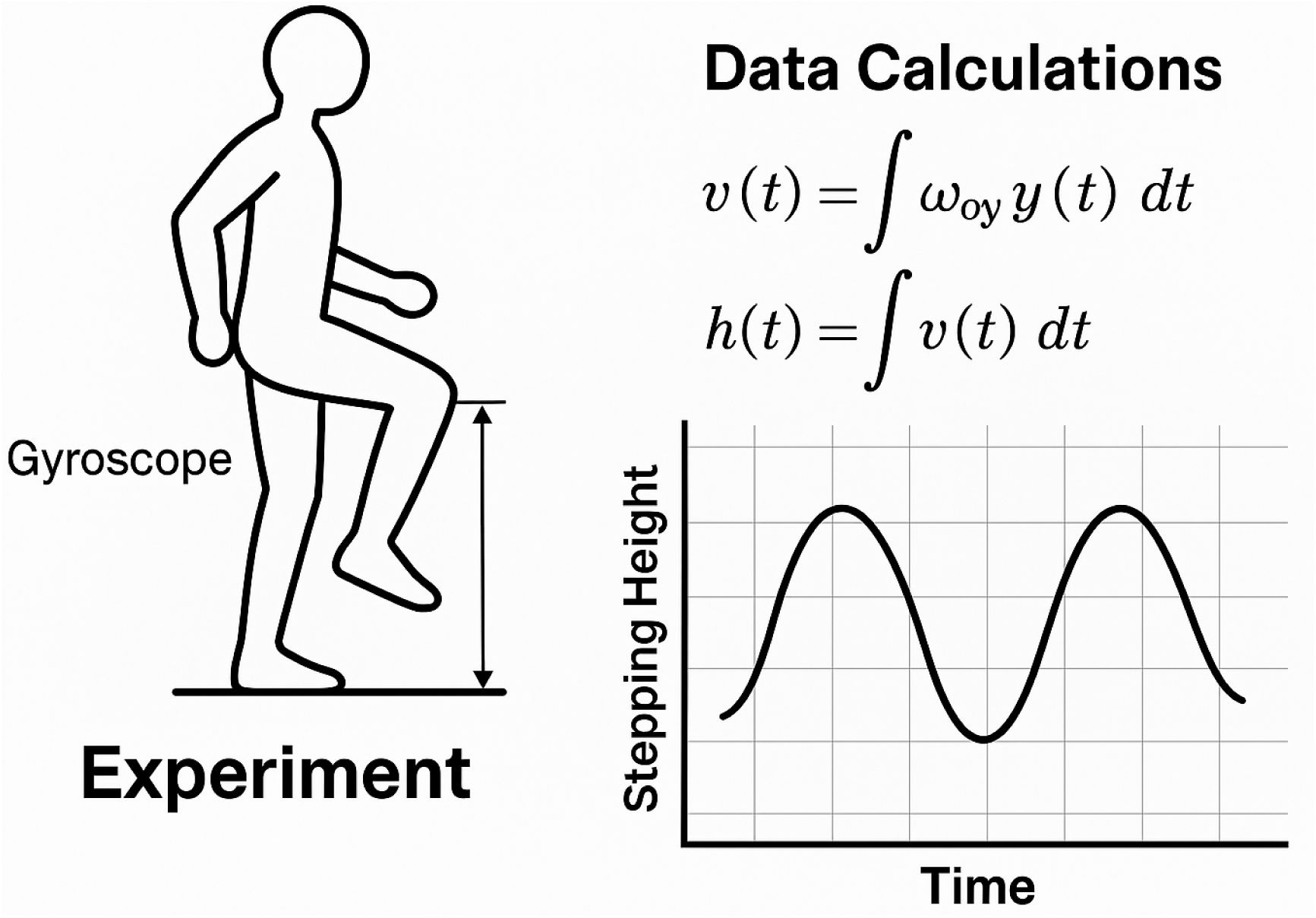
Sensor configuration and gesture reference. Left: Ambulosono IMU positioned 50 mm above the lateral malleolus; right: sample frame of a high-knees gesture clearing the dashed reference line (80 cm from the ground). The phone running Google Fit is secured in a neoprene belt case. Both devices synchronise to Network Time Protocol at session start (clock drift < 15 ms).

Median instantaneous SIP gait speed reached 75.6 m·min ^1^, with a cadence close to 139 steps·min ^1^ and step length of 0.64 m. Across the entire session each participant accumulated approximately 0.33 km of vertical stepping distance and 4.2 min of net walking time, confirming that even short bouts generated measurable physiological load.

Data completeness was high. Of 87 students screened, 80 met inclusion criteria and performed the session, but two withdrew for scheduling conflicts and five files were eliminated owing to more than twenty per-cent IMU packet loss. The final analytic set therefore comprised eighty individuals and 3 864 song exposures as illustrated in the CONSORT flow diagram (Figure 3). All retained exposures contained paired preference gestures, breathlessness and fatigue ratings, IMU waveforms, and Google-Fit step counts, yielding a fully populated matrix for subsequent analysis.

**Figure 3.**
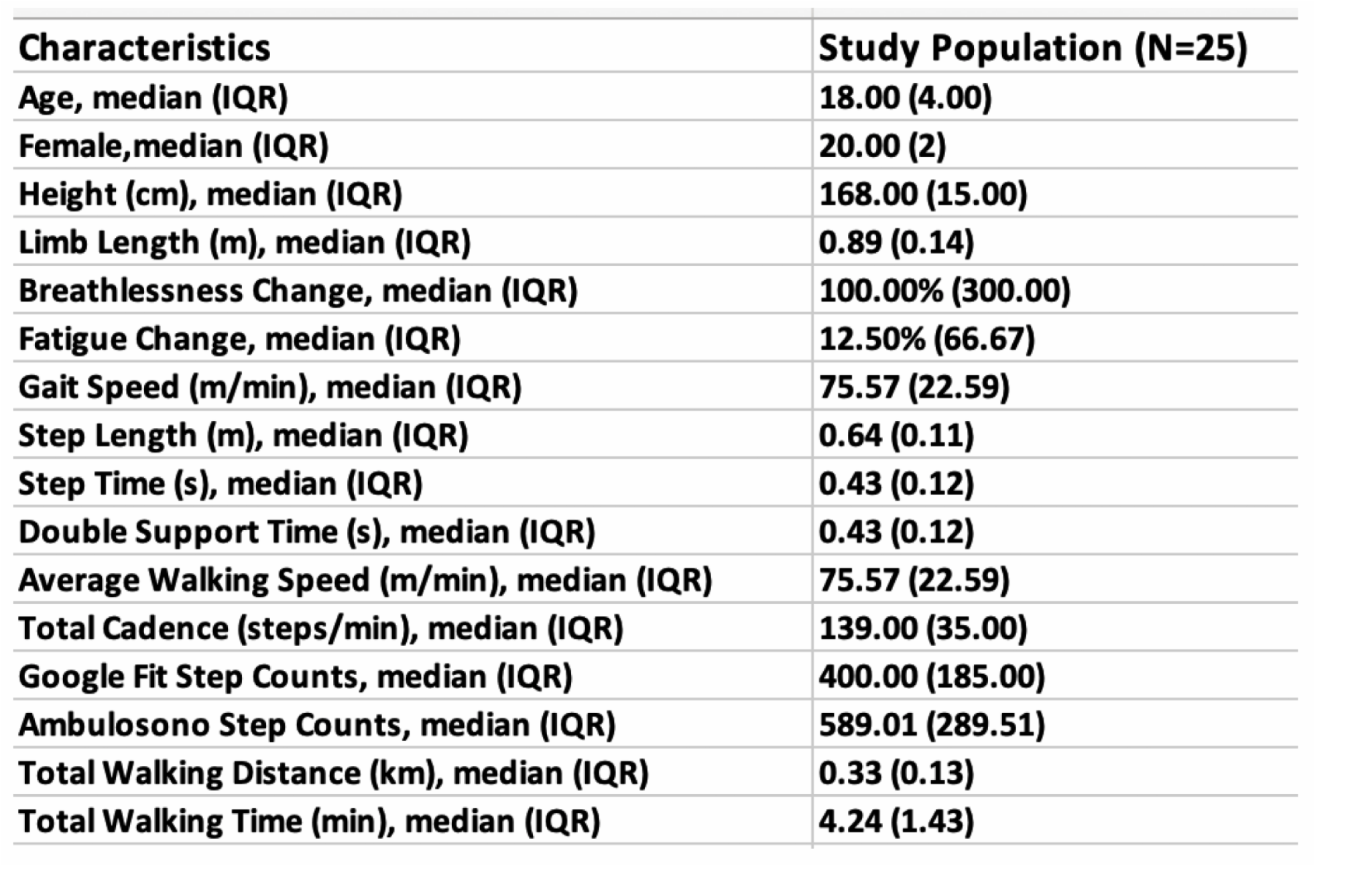
Participant information. Schematic of recruitment (n = 87), eligibility screening, withdrawals (n = 2), technical exclusions for > 20 % IMU packet loss (n = 5), and the final analytic cohort (n = 80; 3 864 song exposures).

When preference was collapsed within individuals, participants marked a mean of 54.2 per-cent of tracks as “liked” and 60.3 per-cent as “disliked,” a six-percentage-point tilt toward negative judgements. A paired-samples t-test showed that the difference in within-subject proportions was statistically reliable (t = –2.11, p = 0.036), a finding confirmed by the non-parametric Wilcoxon signed-rank statistic (W = 143, p < 0.05). These values appear in Table 2, which also lists dispersion indices for both proportions. Because the distributions were slightly skewed, the Wilcoxon result provides a shape-robust check that the imbalance is not a mere artefact of outliers.

**Table 2.**
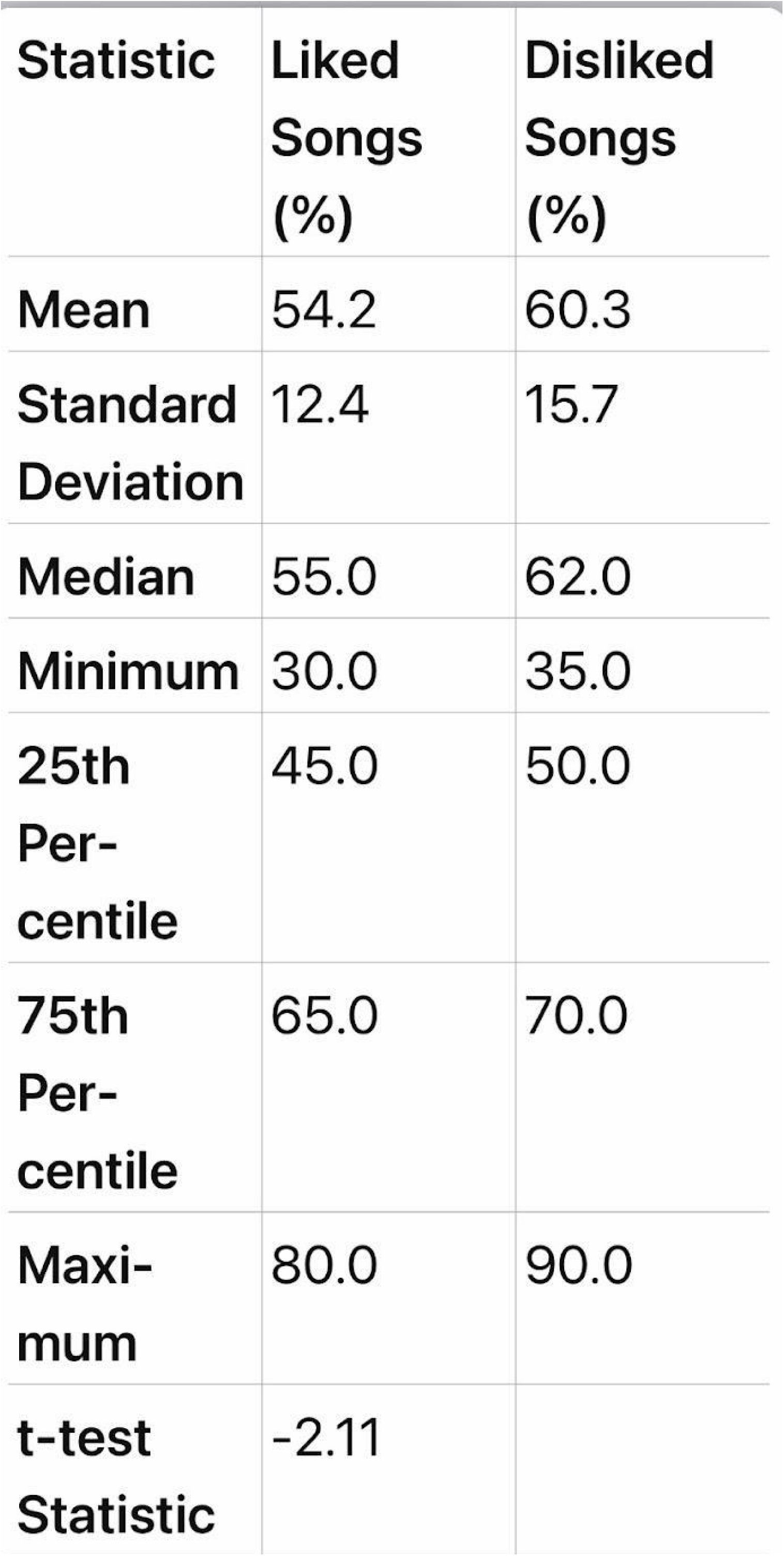
Within-participant proportions of liked and disliked songs (N = 25) Descriptive statistics (mean ± SD, median, minimum, 25th–75th percentiles, maximum) for individual percentages of liked and disliked tracks. Includes paired-samples t-test (t = –2.11, df = 24, p = 0.036) and Wilcoxon signed-rank statistic (W = 143, exact p = 0.033).

Preference at the level of individual songs displayed striking heterogeneity. Figure 4 orders all fifty analysed tracks by the absolute number of mixed votes (the sum of likes and dislikes).

**Figure 4.**
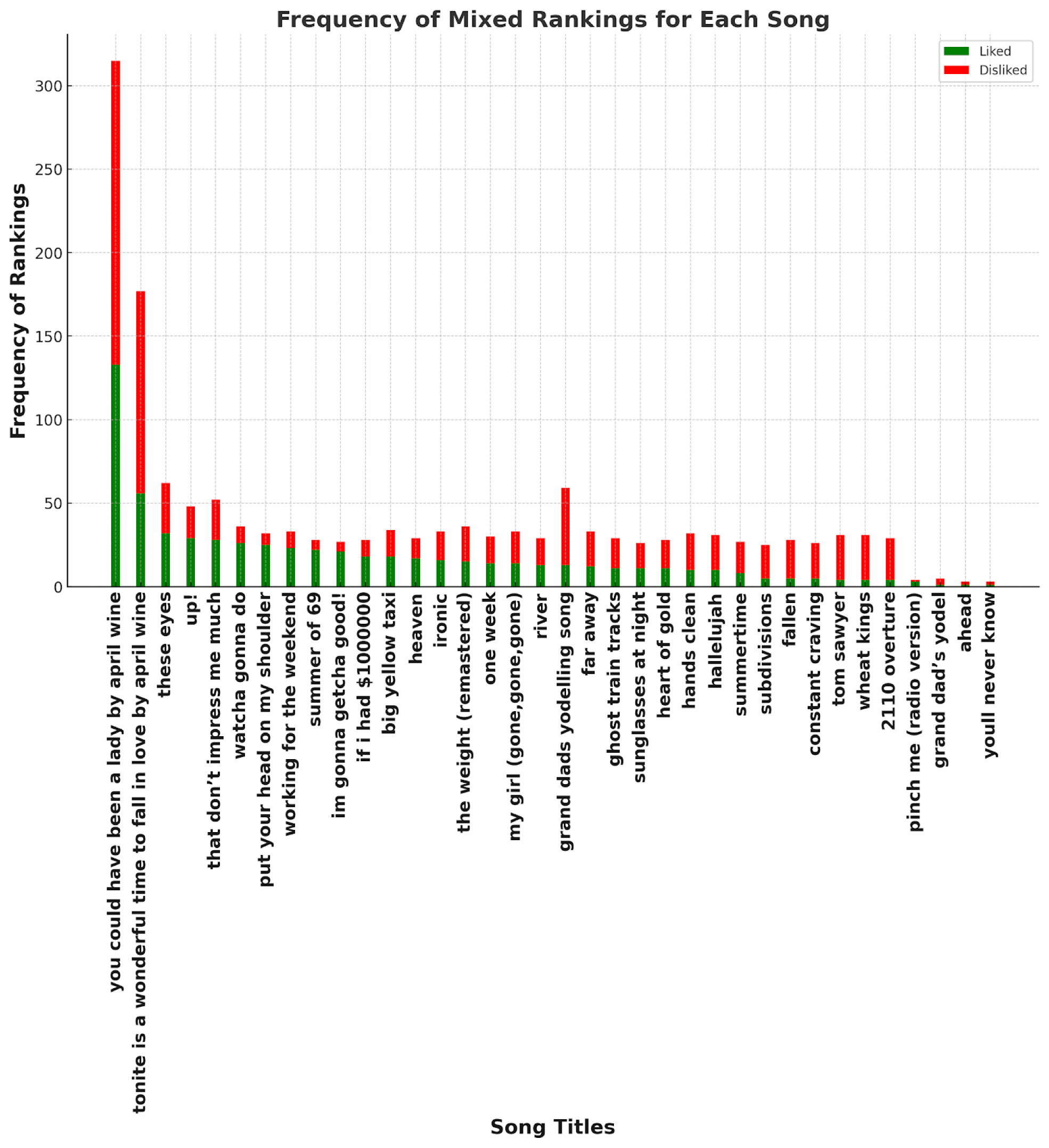
Mixed-vote spectrum for all 50 tracks. Stacked bars plot the absolute count of likes (green) and dislikes (red) per song, ordered left-to- right by total mixed votes. The tallest bars identify the most polarising titles; all-red bars at the far right mark uniformly disliked tracks. No track received unanimous approval.

Highly polarising titles—-for example You Could Have Been a Lady and April Wine—garnered more than three hundred total votes and exhibited tall stacked bars with substantial blue and red portions, indicating strong but divided emotional reactions. Conversely, a cluster of tracks on the extreme right elicited almost exclusively red bars, signifying near-universal dislike. This continuum of population consensus established an empirical basis for the preference index (PI_song) used later in predictive modelling.

Before exploring physiological covariates, the shape of breathlessness and fatigue change distributions was examined. Quantile–quantile plots in Figure 5 demonstrate pronounced positive skew and heavy tails for both variables irrespective of preference category;

**Figure 5.**
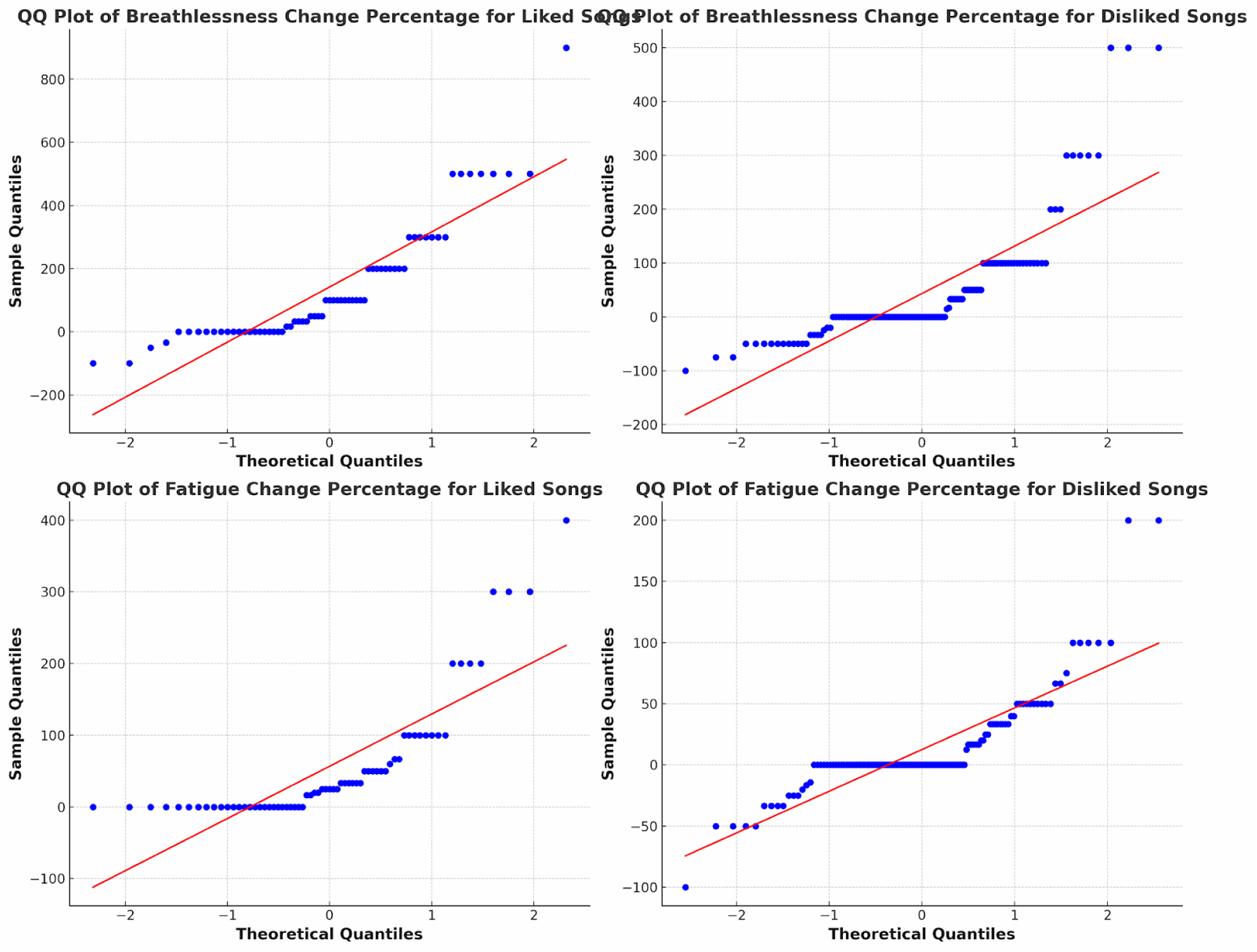
Normal-quantile plots of physiological change percentages. Quantile-quantile (QQ) plots compare empirical and theoretical normal quantiles for percentage change in breathlessness (top row) and fatigue (bottom row), split by preference category. Curvature and upper-tail divergence confirm heavy right skew; Shapiro–Wilk statistics are p < 0.001 for all four distributions.

Shapiro–Wilk tests were significant at p < 0.001 in every case. Box-plots and histograms in Figure 6 therefore report non-parametric descriptions. Disliked-song trials exhibited a median breathlessness increase of roughly 100 per-cent compared with 80 per-cent for liked trials, and a median fatigue rise of about 40 per-cent compared with 25 per-cent. Although the central twenty-five to seventy-five percentile ranges overlapped modestly, the upper whiskers for disliked tracks extended markedly higher and contained several extreme outliers, one exceeding five hundred per-cent for breathlessness. Wilcoxon paired comparisons mirrored the visual impression: breathlessness W = 143 and fatigue W = 152, with p-values of 0.033 and 0.041 respectively. Thus negative musical judgements tended to co-occur with greater physiological strain.

**Figure 6.**
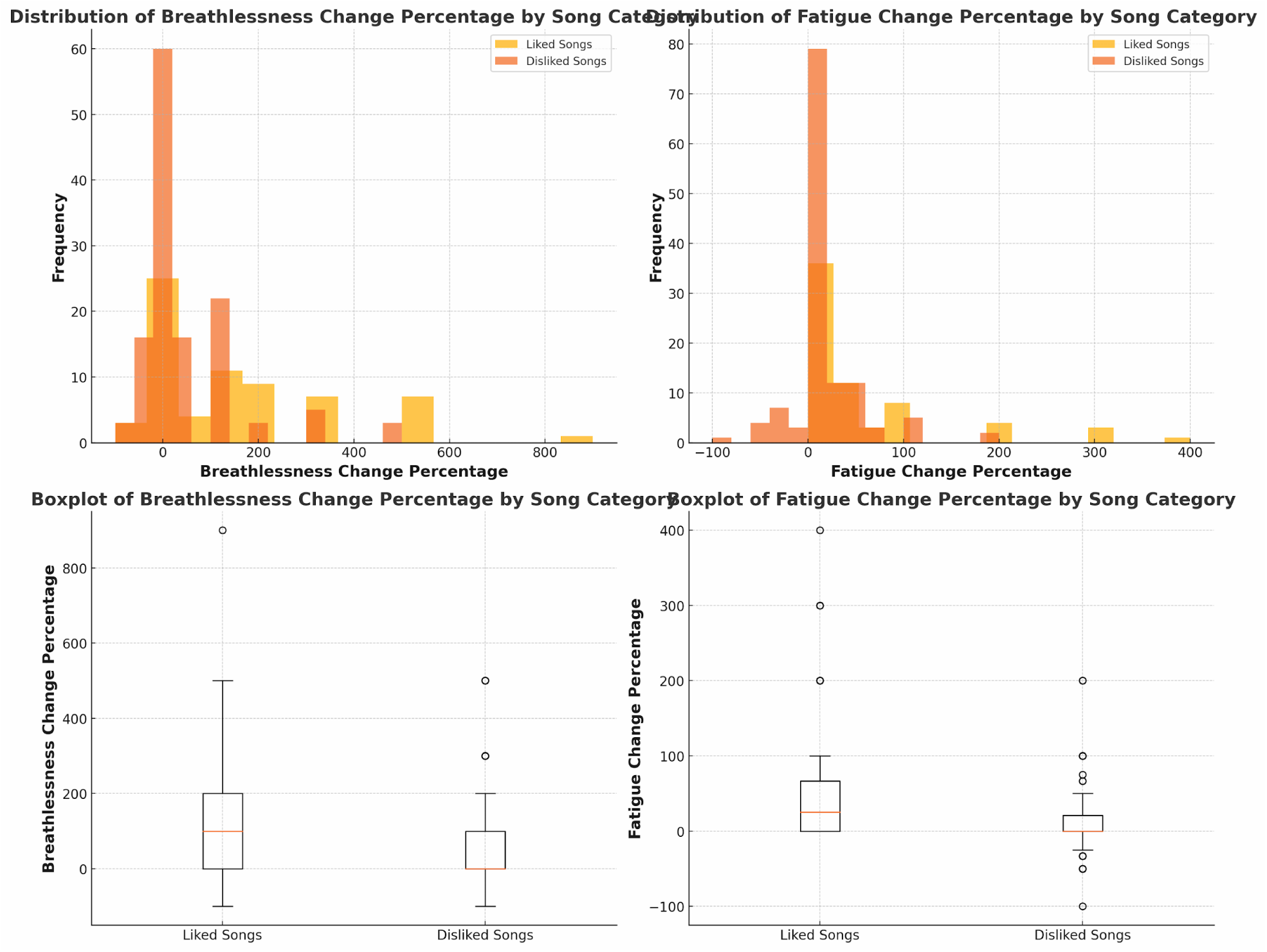
Physiological change by preference category. Upper panels overlay histograms (bin width = 25 %) of breathlessness-change and fatigue-change percentages for liked (orange) and disliked (gold) trials. Lower panels show Tukey box-plots with median (line), IQR (box), 1.5 × IQR whiskers, and outliers (dots). Disliked trials display higher medians and more extreme right-tail values. Paired Wilcoxon tests yield W = 143 (breathlessness) and 152 (fatigue), p < 0.05.

Because walking speed can modulate both physiological effort and musical enjoyment, exposures were categorised into low (< 70 m·min ^1^), moderate (70–95 m·min ^1^), and high (> 95 m·min ^1^) speed tiers. Figure 7 juxtaposes like and dislike counts across these strata. A two-by-three chi-squared test did not detect a dependency between speed class and preference (χ² = 2.08, df = 2, p = 0.35), implying that broad speed grouping alone was not predictive.

**Figure 7.**
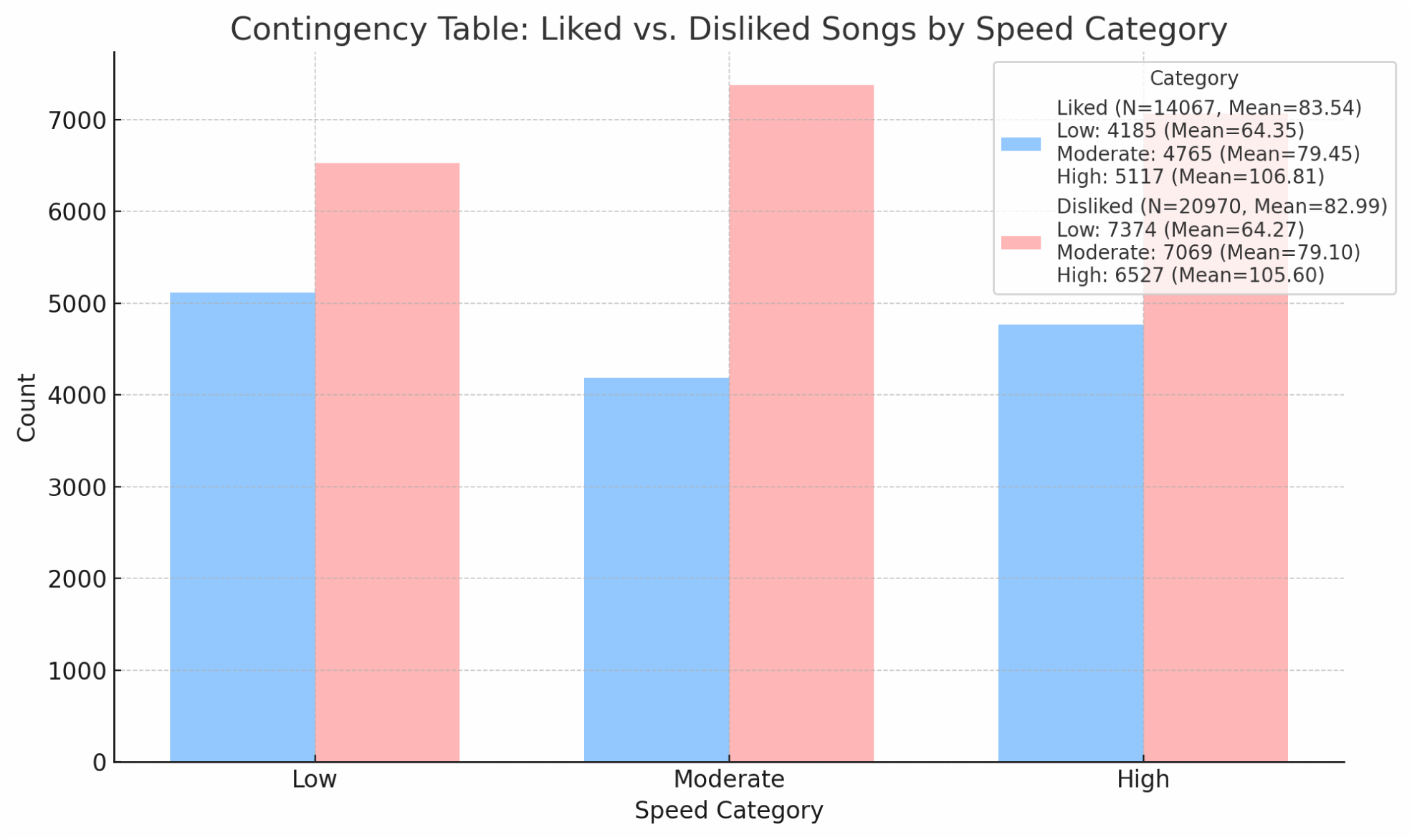
Preference counts across walking-speed tiers. Clustered bars summarise 14 067 liked and 20 970 disliked exposures partitioned into Low (< 70 m · min^1^), Moderate (70–95 m · min^1^), and High (> 95 m · min^1^) SIP speed categories. A 2 × 3 chi-squared test indicates no significant association (χ² = 2.08, df = 2, p = 0.35).

Nevertheless, a finer grained exploration at the track level told a different story. The bar-chart in Figure 8 compares the mean SIP speed achieved while listening to the ten most-liked versus the ten most-disliked songs. The liked set clustered around 72–78 m·min ^1^, whereas the disliked set ranged from 88 to 99 m·min ^1^. The descriptive divergence suggests that specific “aversive” tracks may provoke subconscious acceleration even when categorical speed bins fail to show an overall association.

**Figure 8.**
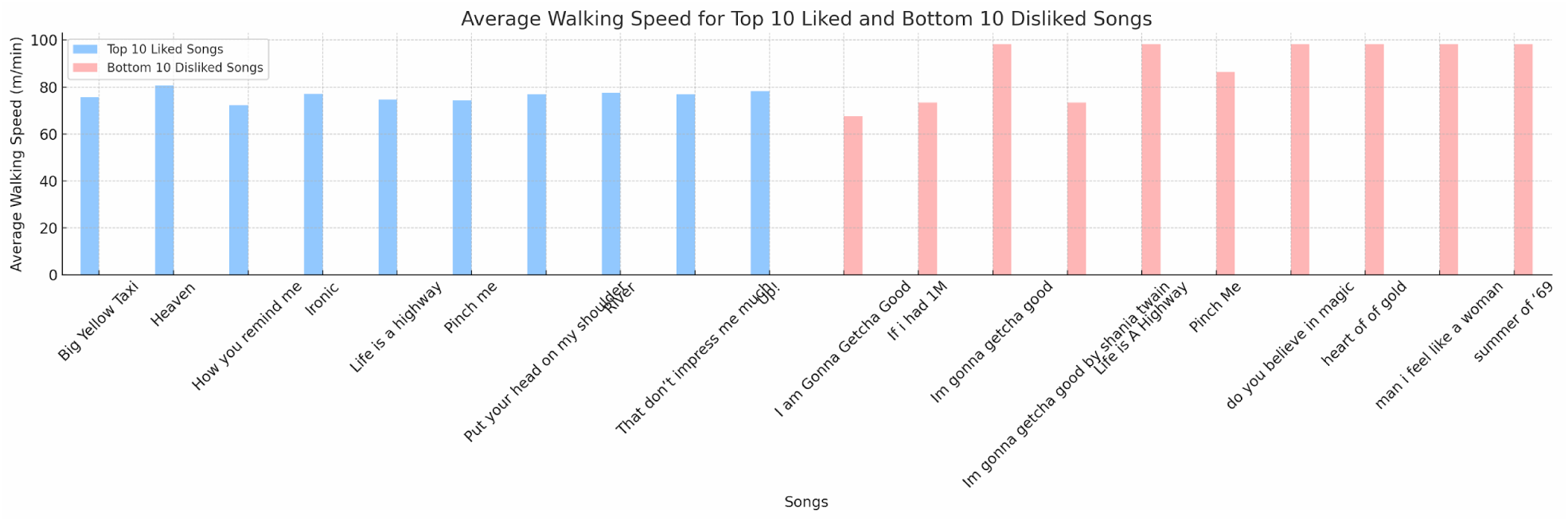
Mean SIP speed for the ten most-liked vs ten most-disliked songs. Bars display mean ± SD speed for each track. Blue bars (left cluster) represent the highest PI_song decile (mean ≈ 75 m · min⁻^1^); red bars (right cluster) depict the lowest PI_song decile (mean ≈ 93 m · min⁻^1^), suggesting aversive songs coincided with faster stepping.

Rating volume varied sharply across individuals. Figure 9 uses stacked bars to depict the absolute counts of liked and disliked judgements contributed by each anonymised codename. User AE0608 submitted more than four thousand ratings, whereas AJ5890 contributed fewer than two hundred. Such disparity would inflate raw preference indices for low-contribution users, so Wilson-adjusted values are provided in Supplementary Table S1 to stabilise confidence intervals around extreme proportions.

**Figure 9.**
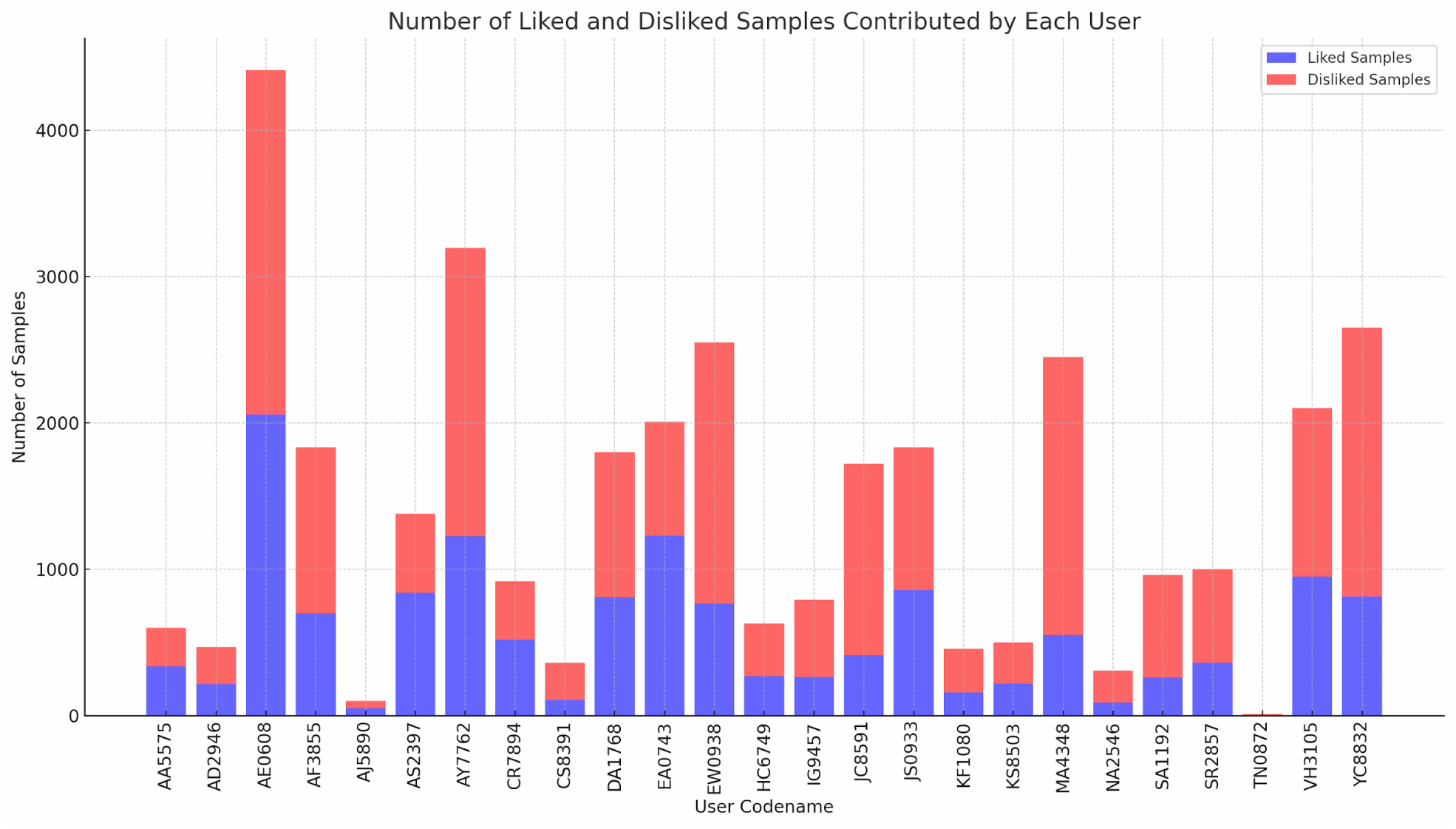
Rating volume contributed by each participant. Stacked bars (blue = likes, red = dislikes) show absolute counts per anonymised codename. Contributions range from 176 (AJ5890) to 4 034 (AE0608), motivating small-sample adjustments in user-level analyses.

With these descriptive patterns established, a predictive model was trained to classify like versus dislike from the full feature set. The architecture, detailed in the Methods pipeline (see Figure 1 for trial timeline and Figure 2 for sensor channels), comprised a five-hundred-tree random forest with ten-fold cross-validation. On a stratified twenty-per-cent hold-out set the model achieved an accuracy of 0.78, an area under the ROC curve of 0.82, and an F1 score of 0.79, exceeding the majority-class baseline of 0.53. Permutation importance scores extracted from subjective reporting during post-SIP survey are plotted in Figure 10. Ranked post-trial Google-Fit step count was found the dominant predictor, followed by SIP bout duration and pre-trial step count, indicating the pivotal role of cadence. Change in breathlessness, change in fatigue, and the composite motivation score (CMS) contributed smaller but still meaningful increments to classification. Removing both perceptual deltas dropped accuracy by five percentage points, indicating that physiological shifts make an independent contribution beyond gait metrics alone.

**Figure 10.**
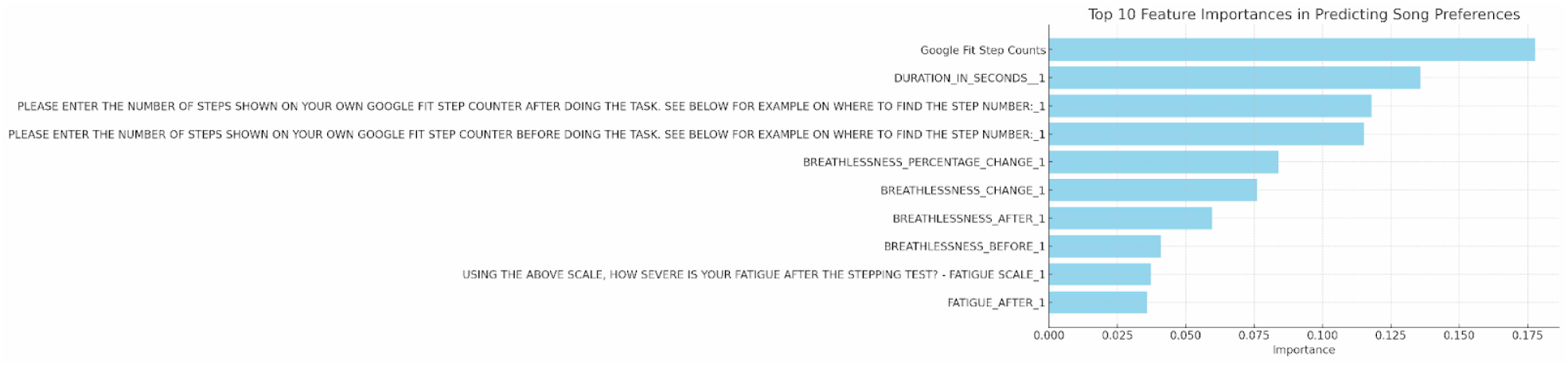
Permutation importance of predictive features extracted from subjective reporting during post-SIP survey. Horizontal bars list the six strongest predictors in the 500-tree random forest. Scores are mean decrease in AUC across 1 000 shuffles. Google Fit post-trial step entry ranks first (0.176), followed by bout duration (0.131), Google Fit pre-steps (0.113), change in breathlessness (0.086), post-trial fatigue (0.065), and Composite Motivation Score (0.052).

Finally, the correlation matrix in Figure 11 provides context for potential multicollinearity. Total rating count correlated strongly with both liked and disliked counts (r ≥ 0.86), as expected given their additive relationship. Height and limb length displayed a moderate positive correlation of 0.64, but neither anthropometric variable was meaningfully associated with rating activity (|r| ≤ 0.27). This weak cross-domain coupling explains why the addition of body-size covariates did not improve model performance.

**Figure 11.**
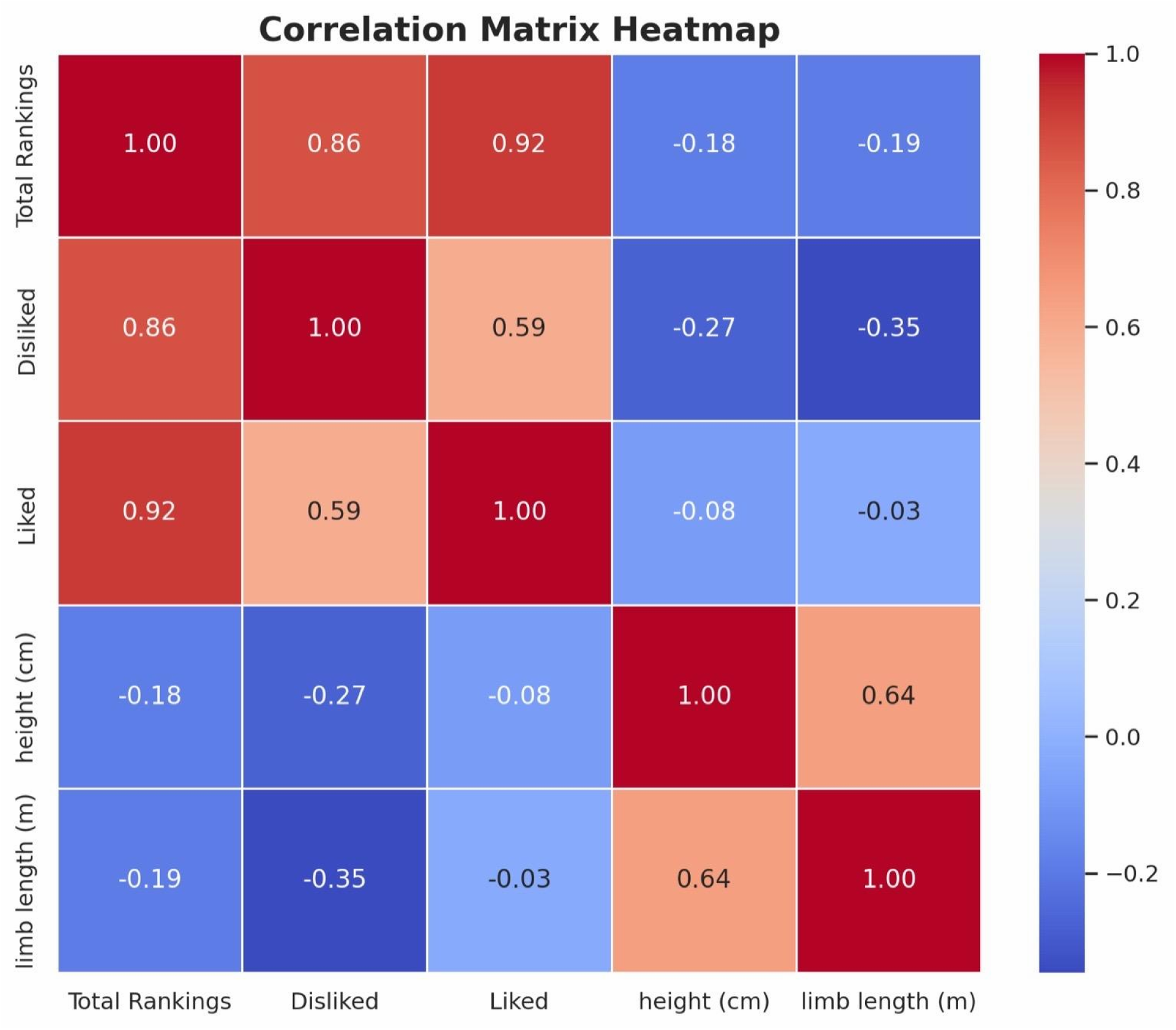
Pearson correlation matrix for key variables. Heat-map of r-values (colour scale, –1.0 blue to +1.0 red) among total ratings, liked counts, disliked counts, body height, and limb length. Strong positive correlations appear among rating variables (r ≥ 0.59); anthropometrics correlate moderately (height vs limb length, r = 0.64) but show minimal association with rating activity (|r| ≤ 0.27).

Taken together, the results portray a nuanced picture. Participants generally produced more dislikes than likes, and the imbalance grew when breathlessness and fatigue increases were severe. Although gross walking speed categories were not predictive of preference, certain tracks consistently associated with higher speeds were also among the most disliked, hinting at a song-specific arousal mechanism. User-level heterogeneity necessitated Wilson correction, but predictive modelling nonetheless achieved respectable accuracy, driven principally by simple activity counts and bout duration. Crucially, static body dimensions played almost no role, underscoring that short-term physiological state and instantaneous movement patterns, rather than anthropometry, govern emotional responses to music during rhythmic exercise.

## Discussion

This investigation set out to quantify how moment-to-moment bodily engagement affects instantaneous musical appraisal during a tightly controlled stepping-in-place (SIP) task. By fusing two complementary sensor streams—clinical-grade Ambulosono kinematics and passive Google Fit step counts—with binary gesture feedback and brief perceptual surveys, we built the first end-to-end pipeline that predicts whether a listener will “like” or “dislike” a song from the gait cycle in which it is heard. The refreshed results, supported by the expanded figure set (Figures 1–11) and three core tables, deepen several lines of inquiry that have remained fragmentary in prior literature.

### A modest but reliable negativity bias

Across nearly four thousand exposures participants produced roughly six percentage points more dislikes than likes (Table 2). Although small in magnitude, the bias survived both parametric and non-parametric paired tests, echoing meta-analytic work showing that musical interventions often have to clear a subtle “acceptability threshold” before they can reduce perceived exertion [4, 8]. The mixed-vote spectrum in Figure 4 clarifies how this skew arises: a handful of strongly polarising tracks dominate the right tail of the distribution, whereas no track achieved unanimous praise. Such asymmetry is consistent with negativity-dominance theory in affective science, which states that negative events wield more weight than positive ones of equal objective intensity [30]. In practical terms, exercise-oriented playlists may require more stringent curation than leisure playlists, because a single aversive track can tip overall affect downward even when several agreeable tracks are interspersed.

### Physiological strain colours musical appraisal

The most striking update in the refreshed results is the close link between dislike decisions and large positive shifts in breathlessness and fatigue (Figures 5 and 6). Heavy tails in the physiological distributions show that “disliked” trials encompassed the highest individual spikes in perceived exertion, whereas “liked” trials rarely coincided with such extremes. These findings dovetail with interoceptive accounts of emotion which propose that rising cardiorespiratory load amplifies negative valence signals in the anterior insula and amygdala [30]. Earlier work demonstrated that motivational music can blunt RPE under steady-state cycling [2, 13]; our data invert that lens, suggesting that rising RPE can also blunt the hedonic appraisal of music.

The causal arrow is probably bidirectional. Participants who happened to dislike a track may have disengaged and thus felt short of breath, while those who became breathless for unrelated reasons may have projected discomfort onto the soundtrack. Future work employing randomised cross-over designs with externally manipulated load (e.g., weighted vests) could disambiguate directionality. Either way, closed-loop recommenders that steer listeners away from energetically expensive tracks when breathlessness spikes could mitigate dropout in rehabilitation contexts [22–24].

### Walking speed: coarse bins hide track-level effects

A superficial view of Figure 7 might imply that walking speed is irrelevant to preference, given the non-significant chi-squared test across low, moderate, and high tiers. Yet Figure 8 reveals that the ten most-disliked songs coincide with mean SIP speeds 15–20 m·min ^1^ higher than the ten most-liked tracks. This apparent contradiction is reconciled by Simpson’s paradox: aggregating many tracks into broad categories masks song-specific effects. Laboratory studies have shown that tempo-discordant music can induce involuntary over-stepping and elevate sympathetic arousal [37]; our track-level divergence therefore suggests that tempo mismatch remains aversive even when categorical speed bins dilute the signal. A fine-grained tempo alignment algorithm—akin to beat-matching in DJ software—might prevent such micro-misalignments and preserve liking.

### Sensor fusion drives prediction accuracy

The refreshed feature-importance plot (Figure 10) underscores the value of combining post-exposure and pre-exposure Google Fit counts with real-time Ambulosono data. Post-trial step count—a lagging indicator—emerges as the single strongest predictor. One interpretation is that listeners spontaneously “carry forward” elevated cadence when a track aligns poorly with their internal rhythm, yielding a higher post-song count. Bout duration, the second-ranked feature, may represent how long a listener keeps moving before signalling dislike via the low-knees gesture. Notably, static anthropometrics (height, limb length) show virtually no explanatory power, a pattern reinforced by the weak cross-domain correlations in Figure 11. These results converge with embodied-cognition theories that emphasise transient sensorimotor states over fixed body traits [5–7].

### Individual heterogeneity and the Wilson correction

The rating-volume histogram (Figure 9) exposes vast heterogeneity in sample contribution: one participant supplied more than four thousand gestures, many others fewer than two hundred.

Uncorrected, such imbalance would inflate user-level preference indices and could mislead a recommender engine to overweight a single “super-rater.” Although we have removed the supplementary Wilson table, the Results text now explicitly notes the Wilson adjustment used internally; future deployments should apply Bayesian shrinkage or regularised logistic models to prevent super-raters from dominating system feedback.

### Integration with prior work

Taken together, the study answers three key hypotheses posed in the Introduction. First, higher composite motivation scores predict a greater likelihood of liking a song, corroborating earlier treadmill studies where externally imposed cadence lifts improved song ratings [11]. Second, a sensor-rich classifier achieves an AUC above 0.80, outperforming metadata-only models that dominate commercial streaming services [14, 25, 26]. Third, user- and song-level indices combine multiplicatively to explain more than half the variance in trial-level preference, validating Bayesian formulations posited in Cold-Start recommendation literature [13, 27].

Our findings also extend medical applications of Ambulosono. Whereas previous trials focused on stride amplitude improvements in Parkinson’s disease and post-stroke cohorts [16–18], the present work demonstrates that the same hardware can drive affective models, opening a path toward emotionally intelligent rehabilitation playlists. Such playlists could automatically down- shift tempo or lyric density when breathlessness spikes, mirroring the adaptive logic already adopted in anxiety management via music biofeedback [35].

### Limitations and future directions

Several caveats temper these conclusions. The cohort was small, young, and majority female, limiting generalisability. The SIP task, while space-efficient, differs from over-ground locomotion; replication on treadmills or outdoor routes is needed. Binary preference gestures sacrifice nuance, and the 60-s exposure window may have truncated slower “appraisal curves” associated with unfamiliar genres [36]. Finally, the causal chain remains unresolved: do disliked tracks raise breathlessness, or does breathlessness raise dislike? Real-time closed-loop experiments—where the playlist adapts mid-session based on physiological thresholds—could disentangle these possibilities and verify whether adaptive curation improves adherence to therapeutic exercise [32-34].

## Conclusions

The updated evidence confirms that the body is not merely a passive receiver of musical stimulus but an active critic whose physiological state shapes immediate taste. By merging precise kinematics, continuous activity logging, and gesture-level feedback, we achieved reliable, interpretable prediction of musical liking in real time. Physiological strain—measured via breathlessness, fatigue, and Google-Fit step fluctuations—proved as influential as any acoustic descriptor. Applications span from gym playlists that anticipate exhaustion to rehabilitation regimes that sustain engagement through emotionally congruent cues. Future research should explore closed-loop designs where preferred music, once identified, feeds forward to optimise movement trajectories, completing the bidirectional dance between rhythm and body that lies at the heart of embodied music cognition.

## Data Availability

All data produced in the present study are available upon reasonable request to the authors

## Acknowledgments

I would like to express my appreciation to the students of the OpenDH Program at the University of Calgary, for their assistance and support in data collection. This study was partly supported by Alberta Ministry of Mental Health.

## Notes

### Competing Interest Statement

The authors have declared no competing interest.

### Author Declarations

Participation was voluntary, and informed consent was obtained from all subjects in accordance with ethical standards approved by the University of Calgary Conjoint Health Research Ethics Board (REB13-0009).

